# The pooled prevalence and associated factors of dropout from maternal continuum of care among mothers in 41 low- and middle-income countries, after the Sustainable Development Goals: a multilevel analysis

**DOI:** 10.1101/2025.11.03.25339360

**Authors:** Mequanent Dessie Bitewa, Milkiyas Solomon, Tadele Derbew Kassie, Mihret Getnet, Ayenew Molla

**Author notes:** **Correspondence** /.

## Abstract

**Background:** This study aimed to assess the pooled proportion of dropouts from the continuum of maternity care (CoC) and its associated factors in LMICs.

**Methods:** A cross-sectional study was conducted based on the data from the Demographic and Health Surveys of 41 low- and middle-income countries after the Sustainable Development Goals. A total of 217,083 (weighted 213,474) women were included. Using STATA v.17.0, a multilevel binary logistic regression analysis was performed to examine the factors associated with dropout from CoC. The adjusted odds ratios with p-values < 0.05 were statistically significant in the final model. The Intra-class Correlation Coefficient, Proportional Change in Variance, and Median Odds Ratio were used to identify the measures of variation for the random effect.

**Results:** The pooled proportion of dropout from CoC was 50% (95% CI: 45% to 56%). Factors associated with higher odds of dropout were multi parity (AOR=1.26, 95% CI: 1.22 to 1.29), grand multi parity (AOR=1.58, 95% CI:1.50 to 1.65), delayed first ANC (AOR=2.87, 95% CI:2.79 to 2.95), WHO regions compared to North Africa/West Asia/Europe region; from SSA (AOR=2.59, 95% CI:2.30 to 2.91), Central Asia (AOR=1.55, 95%CI:1.24 to 1.93), South and Southeast Asia (AOR=2.38, 95% CI: 2.12 to 2.67), Oceania (AOR=2.66, 95% CI:2.20 to 3.23), Latin America and the Caribbean (AOR=9.06 95% CI:7.33 to 11.21), distance from health facility (AOR=1.15, 95% CI:1.12 to 1.19), getting permission (AOR= 1.23, 95% CI:1.19 to 1.28), pregnancy intention (AOR=0.81 95% CI:0.78 to 0.84) and rural residence (AOR=1.52, 95% CI:1.46 to 1.59). While primary education (AOR=0.80, 95% CI:0.77 to 0.83), secondary education (AOR=0.58, 95% CI: 0.55 to 0.60), higher education (AOR=0.44, 95% CI:0.42 to 0.47), middle wealth index (AOR=0.80, 95% CI: 0.78 to 0.83), rich wealth index (AOR=0.67, CI: 0.65 to 0.70), media exposure (AOR=0.73, 95% CI:0.71 to 0.75), women aged between 20 to 34 (AOR=0.80, 95% CI:0.76 to 0.84), and age > 35 years (AOR=0.63, 95% CI:0.59 to 0.67) were associated with lower odds of dropout.

**Conclusions:** Improving women’s education, media exposure, women’s decision-making power, accessibility of health facilities to remote settings, and decreasing teenage pregnancy were found to be factors to reduce dropouts.

## Background

The phrase “continuum of maternity care” describes the delivery of healthcare services by medical professionals to pregnant women throughout pregnancy, labor, and the early postpartum phase, to promote the health and well-being of mothers and their infants. (1). Global maternal health targets for 2030 include achieving at least 90% coverage of four or more prenatal care visits, 90% skilled birth attendance, and 80% postnatal care. National targets aim for 90% coverage of at least four antenatal care visits, skilled birth attendance, and postnatal care (2). A woman is considered to have dropped out of the continuum of maternity care if she fails to complete at least four antenatal care visits (ANC4+), is not attended by a skilled professional during childbirth, or misses the postnatal care (PNC) visit (3). In developing countries, postnatal care within 48 hours of delivery is critical for managing postpartum hemorrhage, a leading cause of maternal mortality (4). The risk of maternal and neonatal deaths is highest during this critical period. The first 48 hours following birth account for a substantial proportion of maternal and newborn deaths worldwide (5, 6). Missing postnatal check-ups in the first two days after birth were responsible for more than half of maternal and neonatal deaths (7, 8). The health and well-being of the mother and baby rely on the standard of maternal care provided during pregnancy, childbirth, and after delivery (9).

Poor labor and delivery outcomes often result from inadequate prenatal care, increasing the risk of complications. Failure to complete the ANC and other components of the maternal continuum of care (CoC) may precipitate the risk of maternal and neonatal death. This is because missed opportunities for early detection and prevention of birth-related complications can have serious consequences (10, 11).

Sustainable Development Goal 3 (SDG 3.1 and 3.2) aims to reduce maternal mortality by at least two-thirds from its 2010 baseline by 2030 in all countries. Between 2000 and 2017, maternal mortality decreased by 2.9%, from 342 to 211 deaths per 100,000 live births. Nevertheless, the present pace of decrease is far less than what is needed to achieve the global goal for 2030. By 2030, countries are expected to lower maternal deaths to less than twice the global target, meaning fewer than 70 maternal deaths per 100,000 live births. (12, 13). Achieving this goal will require substantial effort, particularly in most low-income countries. Globally, avoidable causes related to pregnancy and childbirth claimed the lives of nearly 292,000 women in 2020 (14, 15). Unacceptably high maternal mortality rates persist worldwide, especially in low- and middle-income countries (LMICs), which accounted for approximately 95% of maternal deaths in 2020 (14). Southern Asia and Sub-Saharan Africa alone were responsible for about 86% of all maternal deaths globally (12). Although over 80% of mothers in developing regions are enrolled in antenatal care (ANC), only about half of pregnant women meet the previously recommended number of antenatal visits by the World Health Organization (WHO) (16–19). Furthermore, the transition from antenatal care (ANC) to postnatal care (PNC) is alarmingly low in low-income countries, with completion rates varying widely across nations in low- and middle-income countries (LMICs) (16–20).

Low-income countries have a significantly higher lifetime risk of pregnancy- and birth-related deaths compared to high-income countries. The risk is approximately 1 in 49 in low-income countries versus 1 in 5,300 in high-income countries (14), representing a 19-fold difference. Dropout from the maternity care continuum is directly or indirectly associated with the majority of maternal and neonatal deaths. Missing antenatal care significantly increases the likelihood of home deliveries (21), and mothers who give birth at home are known to have a higher risk of perinatal complications (22). These complications include antepartum, intrapartum, or postpartum hemorrhage, birth trauma, and sepsis, all of which can lead to birth-related deaths (23, 24). Mothers attended by health professionals throughout the perinatal period can save their own lives and those of their babies, thereby reducing maternal and neonatal deaths as well as stillbirths (13, 14, 25). It is also essential to address the barriers that prevent mothers from continuing through the maternity care continuum (26). To achieve reductions in maternal mortality ratios and infant mortality rates, dropout rates from the continuum of maternity care must be decreased. Sustained adoption of these services is critical to improving maternal, neonatal, and child health outcomes in low- and middle-income countries (27). The progress made in reducing maternal deaths so far remains insufficient.

Maternal mortality and morbidity in low- and middle-income countries are largely preventable and are associated with higher dropout rates from maternal healthcare services in these regions (14, 26). To achieve the 2030 Sustainable Development Goal of “Ending preventable maternal, neonatal, and child deaths, low- and middle-income countries must prioritize reducing dropout from the maternal continuum of care. Despite its critical role in decreasing maternal and newborn mortality, there is insufficient data on the extent of dropout from the maternity continuum of care in low- and middle-income regions (28).

The overall rate of dropout from the maternal continuum of care (CoC) in low- and middle-income countries (LMICs), along with the factors that lead to it, is poorly understood. Some previous studies have identified CoC dropout and its contributing factors; however, most of these studies were country-specific, limiting cross-setting comparisons and providing fragmented evidence insufficient to guide global maternal health efforts (29). Few studies have utilized multi-country, standardized data to analyze the proportion and factors associated with dropout across various health system contexts (18). Accordingly, the purpose of this study was to evaluate the pooled proportion of maternal continuum of care dropout and its association with significant variables (maternal age, residence, women’s education, sex of household head, media exposure, wealth index, parity, pregnancy intention, getting permission to seek healthcare, distance to the nearby health facility, and time of initiation of first ANC visit) in the study area.

## Methods and materials

### Data source and study area

A secondary data analysis was conducted using data from the WHO regions encompassing low- and middle-income countries after the Sustainable Development Goals (SDGs). This population-based cross-sectional study utilized data from six WHO regions classified by the Demographic and Health Surveys (DHS) program, covering 41 low- and middle-income countries from 2016 to 2024. A total of 41 countries were selected for analysis. When more than two standard DHS datasets were available after 2016, the most recent dataset was used. Each survey followed a cross-sectional study design, collecting data on independent and outcome variables simultaneously. The data for this study were requested via the DHS program website link (https://dhsprogram.com/data/dataset_admin/index.cfm) once the purpose of the study was explained through the submission of a project summary.

Low- and middle-income countries (LMICs) represent a group of nations, each encountering distinct challenges. The World Bank classifies them into low-income, lower-middle-income, and upper-middle-income categories (30, 31).

### Population

The target population comprised all women aged 15 to 49 in LMICs who had given birth within the two years preceding each DHS survey. The study population included all women aged 15 to 49 in the surveyed areas who had most recently given birth within the two years before each DHS survey and who had initiated antenatal care visits in selected countries.

### Inclusion criteria and exclusion criteria

This study included all women aged 15 to 49 in 41 low- and middle-income countries (LMICs) who had delivered a child within two years before each DHS survey and had initiated antenatal care visits. Women who were pregnant at the time of the surveys, had begun antenatal care, but had subsequently ended their pregnancies, were excluded.

### Sample size determination and sampling procedure

#### Sample size determination

The sample size for our study included all recorded women who gave birth within two years before data collection and had booked antenatal care. Although data on ANC and delivery were documented for up to five years before the survey, PNC data were collected only for the two years preceding the survey (32). Consequently, the sample size of the study may have been reduced. Before applying weights, the study included 217,083 observations; after weighting, the total was 213,474. (Table 1)

**Table 1:** The weighted frequency of women who gave birth before two years of the survey in 41 LMICs DHS surveys from 2016 to 2024, after the SDG.

| S.No. | Countries of LMICs* with different regions | Country code | Survey years | Frequency | Weighted frequency | Percent out of total |
| --- | --- | --- | --- | --- | --- | --- |
| **01** | **Sub-Saharan Africa** |  |  |  |  | **41.96%** |
| 1 | Benin | BJ7 | 2017-18 | 4,619 | 4,659 | 2.36% |
| 2 | Burkina Faso | BF8 | 2021 | 4689 | 4,671 | 2.37% |
| 3 | Burundi | BU7 | 2016-17 | 5,105 | 5,264 | 2.67% |
| 4 | Cameroon | CM7 | 2018 | 3,152 | 3,246 | 1.65% |
| 5 | Cote d'Ivoire | CI8 | 2021 | 3,880 | 3,625 | 1.84% |
| 6 | Democratic Republic of Congo | CD8 | 2023-24 | 7,056 | 6,735 | 3.15% |
| 7 | Ethiopia | ET7 | 2016 | 2,703 | 2,779 | 1.41% |
| 8 | Gabon | GA7 | 2019-21 | 2,206 | 2,214 | 1.12% |
| 9 | Ghana | GH8 | 2022 | 3,702 | 3,345 | 1.57% |
| 10 | Gambia | GM7 | 2019-20 | 3,329 | 3,049 | 1.55% |
| 11 | Guinea | GN7 | 2018 | 2,622 | 2,580 | 1.31% |
| 12 | Kenya | KE8 | 2022 | 7,206 | 6,657 | 3.64% |
| 13 | Lesotho | LS8 | 2023-24 | 1,072 | 959 | 0.45% |
| 14 | Liberia | LB7 | 2019-20 | 2,191 | 2,020 | 1.02% |
| 15 | Madagascar | MD8 | 2021 | 4,291 | 4,296 | 2.18% |
| 16 | Mali | ML7 | 2018 | 2,935 | 3,178 | 1.61% |
| 17 | Mauritania | MR7 | 2019-21 | 3,692 | 3,675 | 1.86% |
| 18 | Mozambique | MZ8 | 2022-23 | 3,205 | 3,178 | 1.49% |
| 19 | Nigeria | NG7 | 2018 | 9,119 | 9,225 | 4.68% |
| 20 | Rwanda | RW7 | 2019-20 | 3,058 | 3,149 | 1.60% |
| 21 | Senegal | SN8 | 2023 | 3,618 | 3,344 | 1.57% |
| 22 | Sierra Leone | SL7 | 2019 | 3,517 | 3,483 | 1.77% |
| 23 | South Africa | ZA7 | 2016 | 1,289 | 1,279 | 0.65% |
| 24 | Tanzania | TZ8 | 2022 | 3761 | 3820 | 1.94% |
| 25 | Uganda | UG7 | 2016 | 5,584 | 5,489 | 2.78% |
| 26 | Zambia | ZM7 | 2018 | 3,878 | 3,840 | 1.95% |
| **02** | **North Africa/ West Asia/Europe** |  |  |  |  | **2.52%** |
| 27 | Albania | AL7 | 2017-18 | 926 | 925 | 0.47% |
| 28 | Jordan | JO8 | 2023 | 2,756 | 2,480 | 1.16% |
| 29 | Turkey | TR7 | 2018 | 936 | 868 | 0.44% |
| **03** | **Central Asia** |  |  |  |  | **1.10%** |
| 30 | Tajikistan | TJ7 | 2023 | 1,466 | 1,493 | 0.70% |
| **04** | **South and Southeast Asia** |  |  |  |  | **52.28%** |
| 31 | Bangladesh | BD7 | 2017-18 | 3,260 | 3,285 | 1.66% |
| 32 | Cambodia | KH8 | 2021-22 | 3,258 | 3,154 | 1.60% |
| 33 | India | IA7 | 2019-21 | 79,516 | 79,216 | 40.15% |
| 34 | Indonesia | ID7 | 2017 | 6,730 | 6,520 | 3.30% |
| 35 | Maldives | MV7 | 2016-17 | 977 | 915 | 0.46% |
| 36 | Nepal | NP8 | 2022 | 1,946 | 1,825 | 0.92% |
| 37 | Pakistan | PK7 | 2017-18 | 3,701 | 3,156 | 1.60% |
| **38** | Philippines | PH8 | 2017 | 2,854 | 2,716 | 1.38% |
| **39** | Timor-Leste | TL7 | 2016 | 2,372 | 2,393 | 1.21% |
| **05** | **Oceania** |  |  |  |  | **1.32%** |
| **40** | Papua New Guinea | PG7 | 2016-18 | 2,691 | 2,595 | 1.32% |
| **06** | **Latin America and the Caribbean** |  |  |  |  | **1.10%** |
| **41** | Haiti | HT7 | 2016-17 | 2,268 | 2,174 | 1.10% |
| **Total** |  |  |  | **217,083** | **213,474** | **100%** |

\* **LMICs: low- and middle-income countries**

#### Sampling procedure

The DHS used a two-stage stratified cluster sampling approach to obtain a nationally representative sample, allowing for comparisons across the country’s regional states. For every survey, the sampling frame was developed using the most recent population and housing census data. The smallest sampling unit, called an enumeration area or cluster, is a geographic region containing several households and functions as a census counting unit. In the first stage, rural and urban areas were stratified, and enumeration areas were selected using probability proportional to size sampling. In the second stage, households within these selected enumeration areas were systematically sampled with equal probability. A detailed household listing was carried out for every chosen enumeration area.

### Study variables

#### Dependent variable

Dropout from a continuum of maternity care: is a dropout from the care of mothers from pregnancy until the postnatal period, including missed WHO-recommended ANC, a birth not attended by skilled personnel, or absence from at least one postnatal check-up within 48 hours after delivery. Participants missed at least one service, which was coded as 1, while those who received all three services were coded as 0 (dropout from CoC = 1, CoC = 0). It was coded 1 if a woman didn’t receive WHO-recommended antenatal care (1–3) and 0 for ANC4+. Among women who received WHO-recommended ANC, some went on to receive skilled birth attendance; some did not. The code was 1 for receiving WHO-recommended ANC but not receiving skilled birth attendance, and 0 for receiving both WHO-recommended ANC and skilled birth attendance. After delivery, some women received postnatal care, and some did not. Finally, a mother who received all services (ANC4+, SBA, and PNC) was coded 0, and a mother who missed at least one service was coded 1.

#### Independent variables

##### Community-level factors

DHS region, place of residence (urban or rural), and distance from a health facility (a big problem or not a big problem).

##### Individual-level factors

Maternal educational attainment (no, primary, secondary, or higher education), mass media exposure (no or there was media exposure), wealth quintile (poor, middle, and rich), parity, getting permission (a big problem or not a big problem), pregnancy intention (wanted or unwanted), age of respondent (<20, 20-34, or >34), timing of first ANC initiation, and sex of household head (male or female) are all factors that were examined for the presence of association with the outcome variable (18, 33–35). (Figure 1)

**Figure 1:**
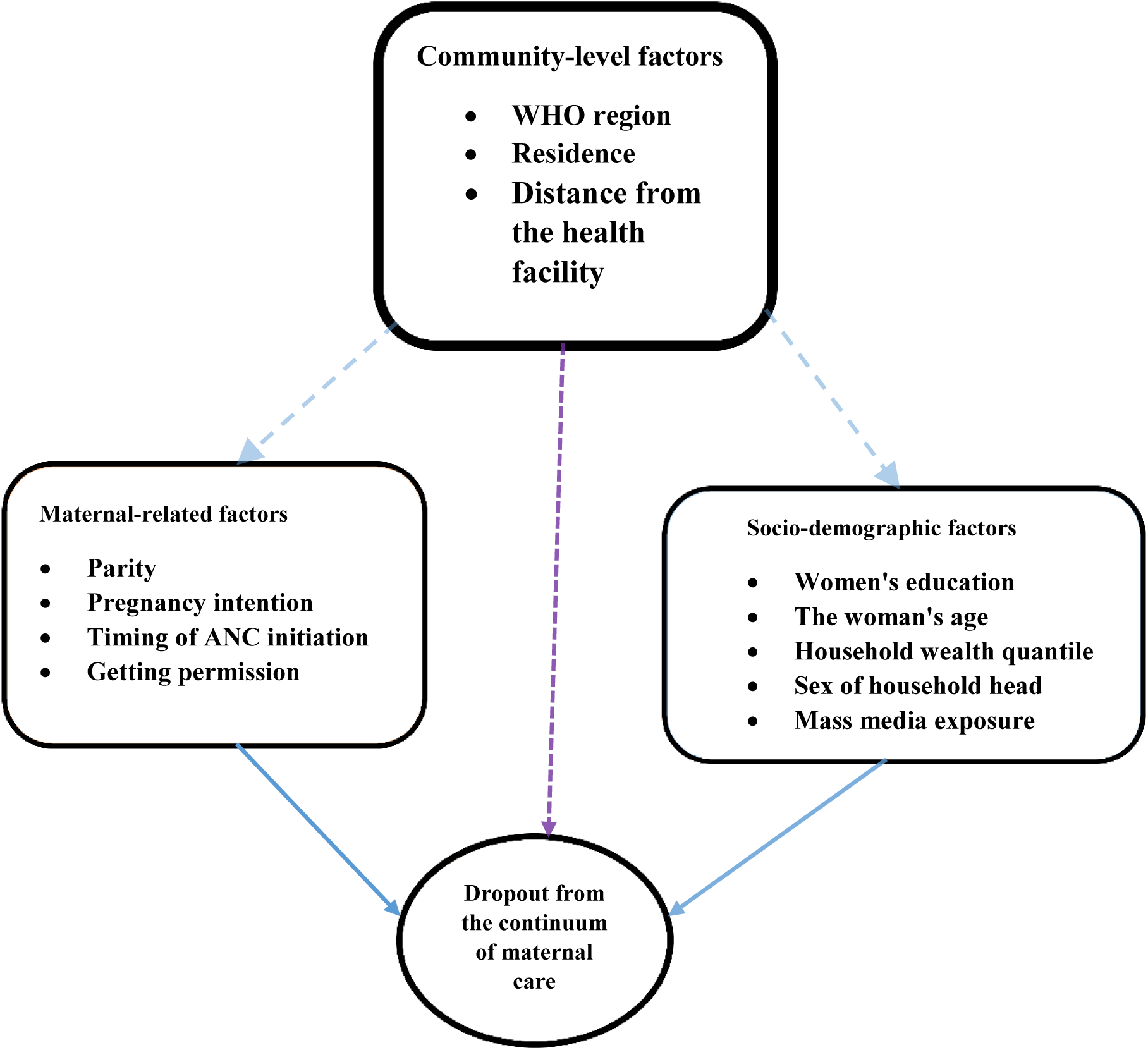
Schematic diagram of the conceptual framework showing potential factors of dropout from the continuum of maternity care in low- and middle-income countries’ DHS from 2016-2024 after SDG

### Data management, data processing, and analysis methods

The study involved a secondary analysis of data obtained from DHS datasets accessible online, collected from 41 low- and middle-income countries (LMICs) following the establishment of the Sustainable Development Goals (SDGs). The data were cleaned, recoded, and analysed using STATA version 17. Following the survey report’s recommendations, the data were weighted to account for sampling probabilities and non-response using the weighting factor to restore the representativeness of the survey and obtain reliable statistical estimates before conducting the analysis. For the independent variables, the assumptions of chi-square and multicollinearity were validated. A bivariable multilevel logistic regression analysis was performed for each independent and dependent variable. Variables with a p-value below 0.2 were then included in the final multivariable multilevel logistic regression analysis. P-values less than 0.05 were considered statistically significant. Adjusted odds ratios (AORs) with 95% Confidence Intervals (CIs) were also calculated. The AORs and their 95% CIs served as measures of association in the full model, which assessed the relationships between the odds of dropping out of the continuum of maternity care and independent factors at both the individual and community levels.

We obtained statistically accurate estimates of the regression coefficients by using clustering information. Therefore, a multilevel analysis was used to obtain the mixed effect (fixed effect for both the individual and community-level predictors and a random effect for the between-cluster variation). The likelihood ratio test was used to determine the significance of the random intercept variation. The intra-class correlation coefficient (ICC), proportional change in variance (PCV), and median odds ratio (MOR) were used to identify the measures of variation for the random effect. 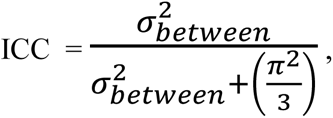 where 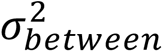 is cluster-level variance in each model (36).

When comparing two individuals from two different randomly selected clusters, the MOR is defined as the median odds ratio between the area at the highest risk and the area at the lowest risk. It was calculated as follows: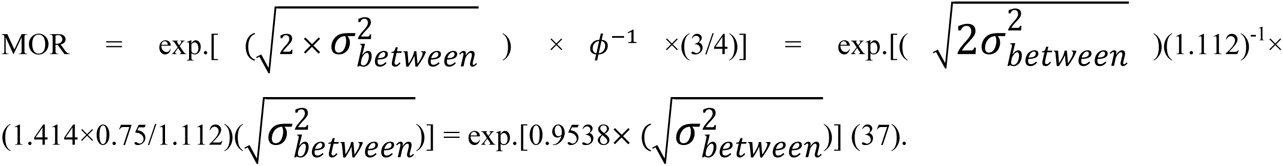

Where *ϕ*^-1^ denotes the inverse of the standard normal cumulative distribution function and 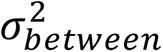 denotes the cluster level variance in each model.

To determine the overall variation at each model attributable to individual and/or community-level characteristics, we employed Proportional Change in Variance (PCV). PCV was calculated using the formula = (VA-VB/VA) ((VA-VB) / VA) × 100, where VA stands for the variance of the empty model and VB for the variance of the model with extra components. Additionally, the candidate model was evaluated using the Deviance Information Criterion (DIC), which is defined as deviance = -2 × log-likelihood.

### Model Building and Comparison

Depending on the continuum of care pathway a woman chooses, she can utilize three services throughout her pregnancy, during birth, and following delivery. She might exit the pathway at any stage, and several factors are considered. Using STATA version 17, four models with variables of interest were fitted to determine which factors contributed to dropout from the continuum of maternal care. Without explanatory variables, the Null Model (empty model) was fitted to test for random variation in the intercept and to estimate the intra-class correlation coefficient (ICC). The ICC is a measure of variation within a cluster. Model I explored the impact of individual characteristics on dropout from maternal care; Model II examined the influence of community-level factors on the outcome; and Model III (the comprehensive model) analyzed the combined effects of individual and community-level characteristics on the outcome simultaneously. The proportional change in variance (PCV), which depicts the change in the community-level variance between the empty model and the succeeding models, and the intra-cluster correlation coefficient (ICC), which is the percentage of variance explained by the higher level, were reported as the measures of variation (community-level variables).

## Results

### Socio-demographic characteristic

Among 213,474 weighted frequencies from 41 LMICs, 89,121 (41.75%) had secondary education, and 97,871 (45.84%) of participants had primary education or less. About 167,155 (78.30%) participants were in the age category of 20-34 years. The 171,169 (80.18%) individuals had planned their most recent pregnancy, and 58.30% (124,458) began their first antenatal care visit within the first three months of pregnancy. More than half, 114,406 (53.59%), of the participants were multiparous mothers. Regarding media exposure, 125,157 (58.63%) had exposure to at least one media during the week (radio, newspapers, or television). Similarly, 89,918 (42.12%) women had poor wealth status. Among participants, about 151,382 (70.91%) and 179,241 (83.96%) did not perceive distance from health facilities and getting permission to seek care as a big problem. Among 213,474 study participants, 141,615 (66.34%) were rural dwellers. Among all study participants, 99,759 (46.73%) and 103,180 (48.33%) women were from Sub-Saharan Africa and South and Southeast Asia, respectively. (Table 2)

**Table 2:**
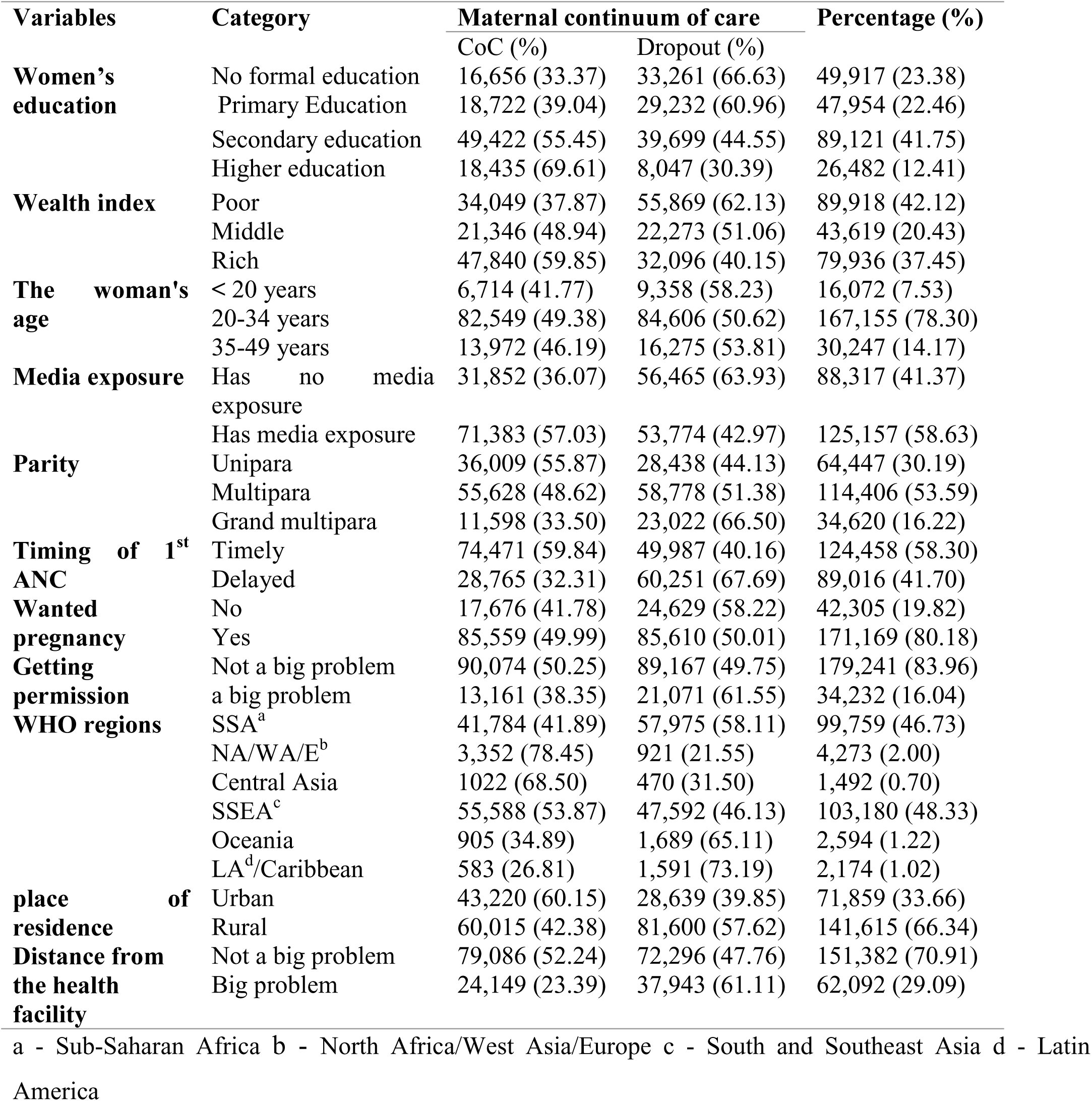
Socio-demographic characteristics of reproductive-age (15-49 years) women who gave birth within two years survey in low- and middle-income countries’ demographic health survey 2016-2024, after SDG.

### Maternal health-related characteristics

The highest dropout rate from previously WHO-recommended antenatal care was observed in Oceania, at 36%. The lowest dropout rate for SBA was in the North Africa/West Asia/Europe region, at approximately 5.30%. The highest SBA dropout rate, at 58%, was observed in Latin America and the Caribbean. For PNC dropouts, the lowest rate was in the North Africa/West Asia/Europe region, around 0.50%, while the highest dropout rate occurred in Latin America and the Caribbean, at 68.50%. The Central Asia region had the lowest PNC dropout rate at 13%. (Figure 2)

**Figure 2:**
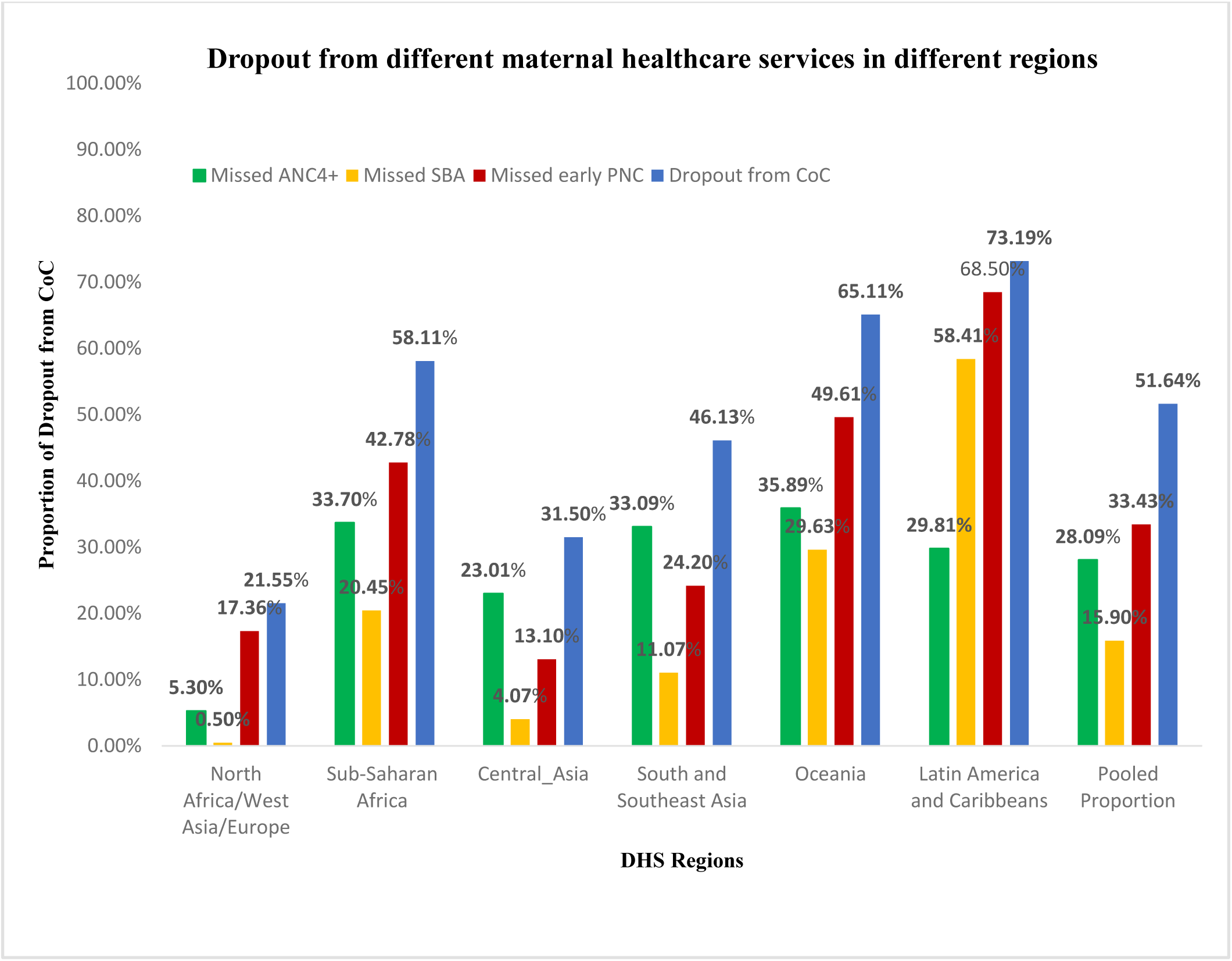
Dropout from different components of maternal health care services by regions of 41 LMICs, as evidenced from the DHS from 2016 to 2024, after the SDG

Maternal continuum of care dropout was higher (73%) in Latin America and the Caribbean region and lower (22%) in the North Africa/West Asia/Europe region. Albania and Turkey had the lowest (19% and 20%) proportion of dropouts, respectively. The proportions of some countries are above the pooled proportion, such as Ethiopia (85%), the Congo Democratic Republic (82%), Guinea (78%), Mozambique (77%), Mauritania (76%), Burundi (74%), Haiti (73%), Madagascar (69%), Timor-Leste (69), Bangladesh (68%), Uganda (66%), Papua New Guinea (65%), Mali (65%), Nigeria (64%), Rwanda (63%), Tanzania (61%), Benin (57%), and Cote d’Ivoire (57%). Lower dropout was found in Albania (19%), Jordan (21%), Indonesia (24%), Ghana (25%), the Maldives (25%), Turkey (27%), Cambodia (27%), Lesotho (28%), and Sierra Leone (29%). (Figure 3)

**Figure 3:**
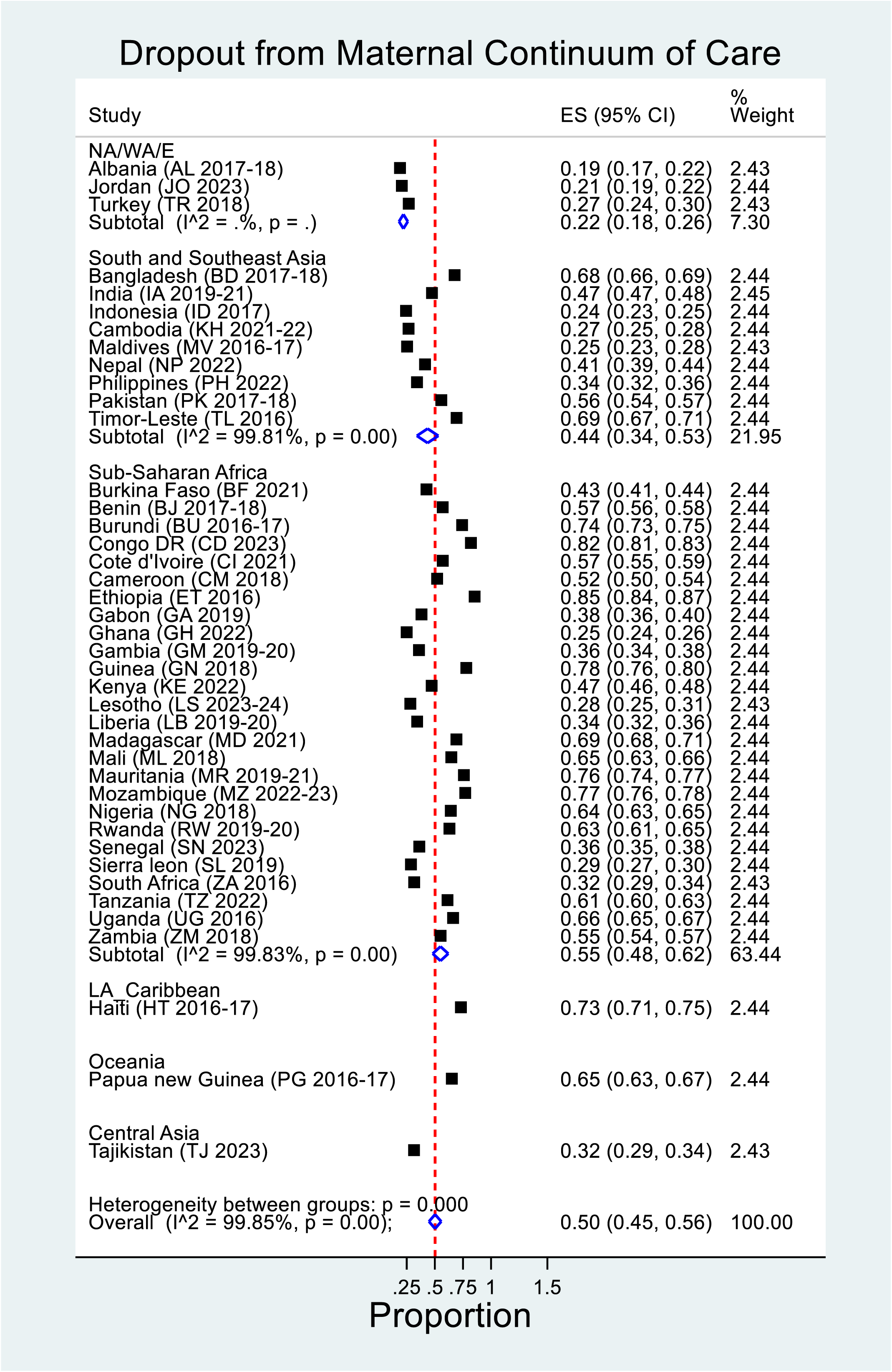
The pooled proportion of dropout from maternal continuum of care by region and country in 41 LMICs, as evidenced from the DHS from 2016 to 2024, after SDG

### Factors associated with a dropout from a continuum of maternal care

In the multilevel analysis, all variables from both individual- and community-level factors with a p-value less than 0.2 in the bivariable analysis were included. In the final model, individual- and community-level factors were analyzed simultaneously. The individual-level factors significantly associated with dropout from the maternal continuum of care (p < 0.05) included women’s education, age, parity, timing of first antenatal care (ANC) initiation, media exposure, getting permission to seek healthcare, and wealth index. At the community level, factors such as the WHO region, residential location, and the distance to a health facility were statistically significant predictors of discontinuation from the maternal continuum of care.

Women with primary, secondary, and higher education were 20% (AOR=0.80, 95% CI: 0.77 to 0.83), 42% (AOR=0.58, 95% CI: 0.55 to 0.60), and 56% (AOR=0.44, 95% CI: 0.42 to 0.47) less likely to drop out of maternal care compared to women with no formal education, respectively. Women with middle and rich wealth indices had a 20% (AOR=0.80, CI: 0.78 to 0.83) and 33% (AOR=0.67, CI: 0.65 to 0.70) lower likelihood of dropping out of the continuum of care compared to women with a poor wealth index, respectively. Among people exposed to media, the odds of dropping out from the continuum of care (CoC) were reduced by 27% (AOR = 0.73, 95% CI: 0.71 to 0.75). The odds of dropout from CoC decreased by 20% (AOR = 0.80, 95% CI: 0.76 to 0.84) among women aged 20 to 34 years and by 37% (AOR = 0.63, 95% CI: 0.59–0.67) among women aged 35 years or older, compared to those under 20 years of age. Similarly, the odds of dropping out were 1.26 times higher (AOR = 1.26, 95% CI: 1.22 to 1.29) among multiparous women and increased by 58% (AOR = 1.58, 95% CI: 1.50 to 1.65) among grand multiparous women compared to uniparous women.

In addition, compared to women who began antenatal care (ANC) visits before three months of gestation, those who delayed initiating ANC visits were 2.87 times more likely to drop out of the continuum of care (CoC) (AOR = 2.87, 95% CI: 2.79 to 2.95). Furthermore, women who perceived obtaining permission to seek healthcare as a significant barrier had a 23% higher likelihood of dropping out before completing maternal care (AOR = 1.23, 95% CI: 1.19 to 1.28). Conversely, the odds of dropping out of the maternal continuum of care decreased by 19% among women who planned their pregnancies (AOR = 0.81, 95% CI: 0.78 to 0.84) compared to those who did not.

At the community level, factors such as WHO region, place of residence, and distance from home to the nearest health facility are significantly associated with the outcome of interest in this study. In the Sub-Saharan Africa (SSA) and Central Asia regions, the odds of dropout from the continuum of care (CoC) increased by 2.59 times (AOR = 2.59, 95% CI: 2.30 to 2.91) and by 55% (AOR = 1.55, 95% CI: 1.24 to 1.93), respectively, compared to the North Africa/West Asia/Europe region. Similarly, the odds increased to 2.38 times (AOR = 2.38, 95% CI: 2.12 to 2.67) in South and Southeast Asia compared to the North Africa/West Asia/Europe region. The risk of dropout increased by 2.66 times (AOR = 2.66, 95% CI: 2.20 to 3.23) in Oceania and by 9 times (AOR = 9.06, 95% CI: 7.33 to 11.21) in Latin America and the Caribbean. Compared to women who did not perceive distance to a health facility as a major problem, those who perceived it as a significant barrier had 15% higher odds of dropping out before completing maternal care (AOR = 1.15, 95% CI: 1.12 to 1.19). Furthermore, women living in rural areas were 1.52 times more likely to discontinue the CoC compared to those living in urban areas (AOR = 1.52, 95% CI: 1.46 to 1.59).

### Random Effects and Model Comparison

The empty model’s intra-class correlation coefficient (ICC) revealed that variations between cluster areas accounted for 42.02% (ICC = 0.4201595) of the overall variation in dropout from the maternal continuum of care, with individual differences accounting for the remaining 57.98% of the variability. In the final model (Model III), the total variation in dropout at the cluster level was reduced to 33.03% (ICC = 0.3302943), which may be attributable to other unobserved community-level factors. Model fitness was compared using deviance, and the model with the lowest deviance (Model III) was considered the best-fitting model. The final model (Model III) had the lowest deviance (1,058.82) compared to the null model (3,228.76) and was used to identify factors significantly associated with dropout from the maternal continuum of care among mothers who gave birth within two years of the survey and had ANC visits in 41 LMICs. Similarly, Model III had the lowest median odds ratio (MOR) value (3.37), indicating that the effects of community heterogeneity were lower compared to the null model’s MOR of 4.36. The Proportional Change in Variance (PCV) shows that both individual- and community-level factors included in the full model explained 31.94% of the variation in dropout from the maternal continuum of care across communities. Multicollinearity among predictor variables was tested using the variance inflation factor (VIF) and was found to be less than 10 (mean VIF = 1.23) in the final model, indicating no multicollinearity. (Table 3)

**Table 3:**
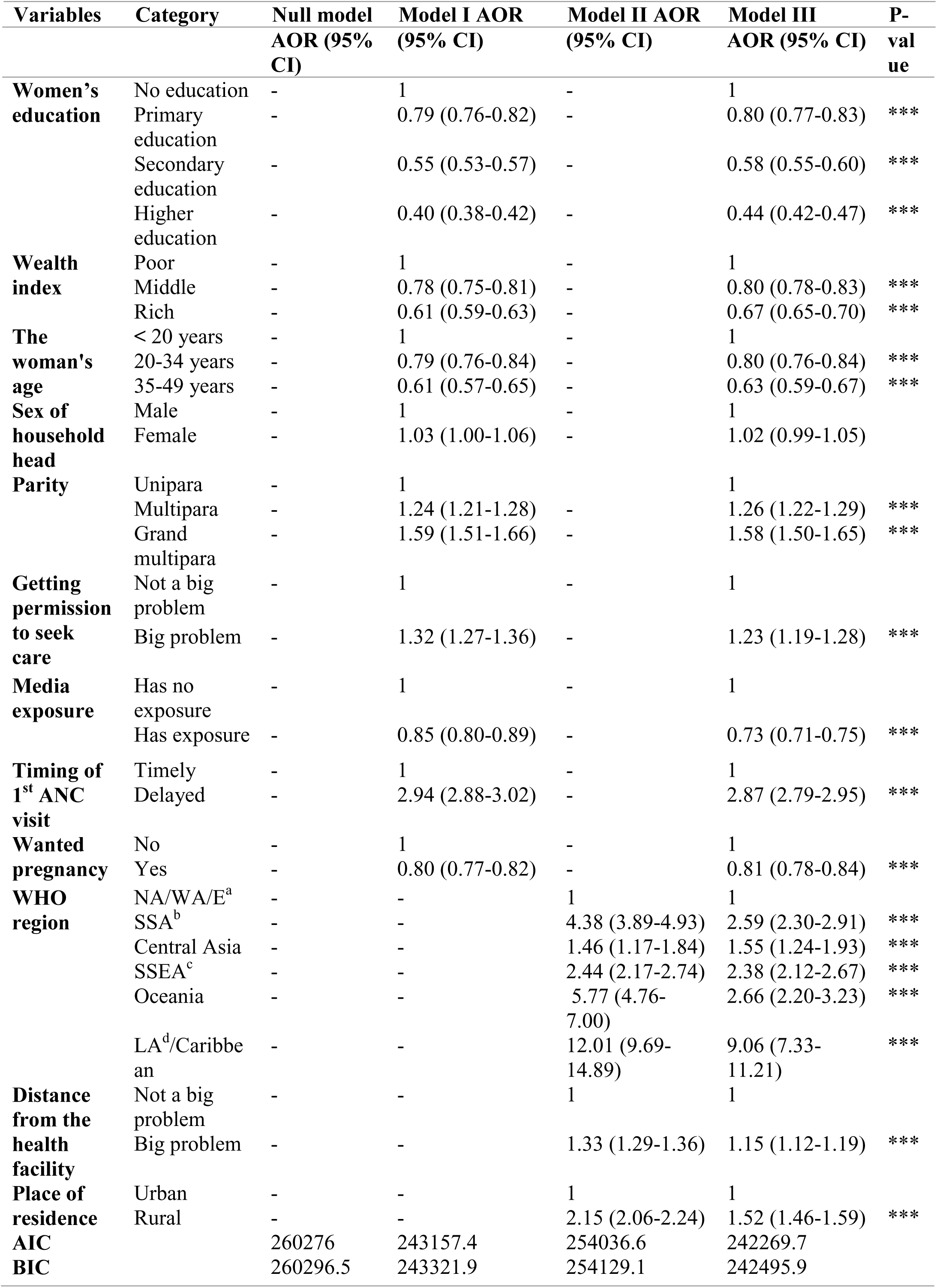
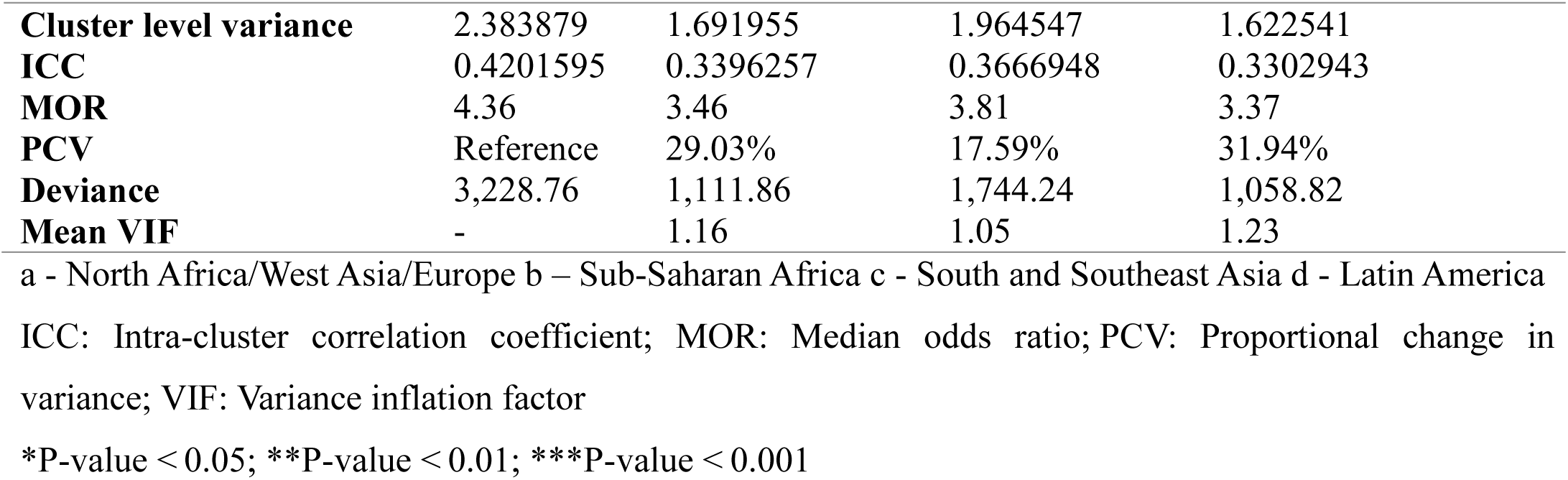
Multilevel analysis of factors associated with dropout from the continuum of care among reproductive women 15 to 49 years in Low- and Middle-Income Countries: based on DHS from 2016 to 2024 after SDG.

| Variables | Category | Null model | Model I AOR | Model II AOR | Model III | P- |
| --- | --- | --- | --- | --- | --- | --- |

|  |  | AOR (95% CI) | (95% CI) | (95% CI) | AOR (95% CI) | value |
| --- | --- | --- | --- | --- | --- | --- |
| **Women's education** | No education | - | 1 | - | 1 |  |
|  | Primary education | - | 0.79 (0.76-0.82) | - | 0.80 (0.77-0.83) | *** |
|  | Secondary education | - | 0.55 (0.53-0.57) | - | 0.58 (0.55-0.60) | *** |
|  | Higher education | - | 0.40 (0.38-0.42) | - | 0.44 (0.42-0.47) | *** |
| **Wealth index** | Poor | - | 1 | - | 1 |  |
|  | Middle | - | 0.78 (0.75-0.81) | - | 0.80 (0.78-0.83) | *** |
|  | Rich | - | 0.61 (0.59-0.63) | - | 0.67 (0.65-0.70) | *** |
| **The woman's age** | < 20 years | - | 1 | - | 1 |  |
|  | 20-34 years | - | 0.79 (0.76-0.84) | - | 0.80 (0.76-0.84) | *** |
|  | 35-49 years | - | 0.61 (0.57-0.65) | - | 0.63 (0.59-0.67) | *** |
| **Sex of household head** | Male | - | 1 | - | 1 |  |
|  | Female | - | 1.03 (1.00-1.06) | - | 1.02 (0.99-1.05) |  |
| **Parity** | Unipara | - | 1 | - | 1 |  |
|  | Multipara | - | 1.24 (1.21-1.28) | - | 1.26 (1.22-1.29) | *** |
|  | Grand multipara | - | 1.59 (1.51-1.66) | - | 1.58 (1.50-1.65) | *** |
| **Getting permission to seek care** | Not a big problem | - | 1 | - | 1 |  |
|  | Big problem | - | 1.32 (1.27-1.36) | - | 1.23 (1.19-1.28) | *** |
| **Media exposure** | Has no exposure | - | 1 | - | 1 |  |
|  | Has exposure | - | 0.85 (0.80-0.89) | - | 0.73 (0.71-0.75) | *** |
| **Timing of 1<sup>st</sup> ANC visit** | Timely | - | 1 | - | 1 |  |
|  | Delayed | - | 2.94 (2.88-3.02) | - | 2.87 (2.79-2.95) | *** |
| **Wanted pregnancy** | No | - | 1 | - | 1 |  |
|  | Yes | - | 0.80 (0.77-0.82) | - | 0.81 (0.78-0.84) | *** |
| **WHO region** | NA/WA/E<sup>a</sup> | - | - | 1 | 1 |  |
|  | SSA<sup>b</sup> | - | - | 4.38 (3.89-4.93) | 2.59 (2.30-2.91) | *** |
|  | Central Asia | - | - | 1.46 (1.17-1.84) | 1.55 (1.24-1.93) | *** |
|  | SSEA<sup>c</sup> | - | - | 2.44 (2.17-2.74) | 2.38 (2.12-2.67) | *** |
|  | Oceania | - | - | 5.77 (4.76-7.00) | 2.66 (2.20-3.23) | *** |
|  | LA<sup>d</sup>/Caribbean | - | - | 12.01 (9.69-14.89) | 9.06 (7.33-11.21) | *** |
| **Distance from the health facility** | Not a big problem | - | - | 1 | 1 |  |
|  | Big problem | - | - | 1.33 (1.29-1.36) | 1.15 (1.12-1.19) | *** |
| **Place of residence** | Urban | - | - | 1 | 1 |  |
|  | Rural | - | - | 2.15 (2.06-2.24) | 1.52 (1.46-1.59) | *** |
| **AIC** |  | 260276 | 243157.4 | 254036.6 | 242269.7 |  |
| **BIC** |  | 260296.5 | 243321.9 | 254129.1 | 242495.9 |  |

|  |  |  |  |  |
| --- | --- | --- | --- | --- |
| **Cluster level variance** | 2.383879 | 1.691955 | 1.964547 | 1.622541 |
| **ICC** | 0.4201595 | 0.3396257 | 0.3666948 | 0.3302943 |
| **MOR** | 4.36 | 3.46 | 3.81 | 3.37 |
| **PCV** | Reference | 29.03% | 17.59% | 31.94% |
| **Deviance** | 3,228.76 | 1,111.86 | 1,744.24 | 1,058.82 |
| **Mean VIF** | - | 1.16 | 1.05 | 1.23 |

a - North Africa/West Asia/Europe b – Sub-Saharan Africa c - South and Southeast Asia d - Latin America
ICC: Intra-cluster correlation coefficient; MOR: Median odds ratio; PCV: Proportional change in variance; VIF: Variance inflation factor
\*P-value < 0.05; \*\*P-value < 0.01; \*\*\*P-value < 0.001

## Discussion

The current study aimed to determine the pooled proportion of women aged 15-49 in low- and middle-income countries who dropped out of maternal care and the factors influencing it, focusing on women who gave birth before two years of the survey and initiated antenatal care after the adoption of the Sustainable Development Goals.

After weighting, the pooled proportion of dropouts from maternal continuum care among reproductive women in 41 low- and middle-income countries after SDG was found to be 0.50 (95% CI: 0.45 to 0.56). The pooled proportion of dropouts was found to be high. Regionally, that was higher in Latin America and the Caribbean region, with 73% (95% CI: 71% to 75%). A low proportion of 22% (95% CI: 18% to 26%) of dropouts occurred in North Africa/West Asia/Europe. The findings underscore substantial regional disparities. This proportion was 3 times less than that of Latin America and the Caribbean. This high discrepancy might be due to only one country (Haiti) being included in the Latin America and the Caribbean region. A single country’s proportion of dropouts may not represent that of the region. Figure 2 revealed that in North Africa/West Asia/Europe, more than 95% and almost all (99.50%) of participants attended the previous WHO-recommended ANC and institutional delivery, respectively. It could diminish the dropout. Furthermore, in North Africa/West Asia/Europe, countries are internally stable and could achieve millennium development goals. For example, Turkey was among the ten countries that successfully met the Millennium Development Goals by significantly reducing maternal continuum dropout rates. Similarly, Albania outperformed other low- and middle-income countries (LMICs) in this area. Nations in this region have demonstrated notable progress toward achieving the Sustainable Development Goals (SDGs) (38, 39). The proportions of dropouts were lowest in Albania (19%), Jordan (20%), and Turkey (27%). In contrast, there were the highest dropouts in SSA, Latin America and the Caribbean, and South and Southeast Asian countries. The proportions of dropouts in several countries were significantly greater than the pooled average. These might be due to different justifications, which might be due to the low-quality service, socio-cultural and economic issues, limited awareness and low health literacy, lack of respectful treatment, or other possible causes that might contribute to the higher dropout (40–43).

Ethiopia, the Democratic Republic of Congo, Ghana, Mauritania, Burundi, Madagascar, Timor-Leste, Bangladesh, Uganda, Papua New Guinea, Mali, Nigeria, Rwanda, Tanzania, Benin, Côte d’Ivoire, and Haiti are classified as least developed countries (44). These countries experience higher dropout rates, which may result from various health system factors such as limited health infrastructure, shortages of skilled providers, long travel distances to health facilities, and financial constraints (45, 46). In countries like Timor-Leste (47) and Ethiopia (48), health system challenges, including limited provider capacity, fragmented health financing, lack of medical supplies and equipment, and poor working and living conditions, disrupt health service delivery. These bottlenecks likely contribute to the higher prevalence of continuum of care (CoC) dropout. Other contributing factors in many resource-limited countries include low socioeconomic status, limited education, and poor quality of services (49). In sub-Saharan Africa (SSA), healthcare service was not perceived as a top priority over the past decade (50).

The findings of this study were consistent with those of a study conducted in Egypt, which reported a rate of 49.6% (35). However, the study in Egypt was a single cross-sectional study, making direct comparison with the current study, which utilized a large aggregated dataset, challenging. Another difference is that the previous study analyzed data spanning five years from birth, whereas the current study included mothers who gave birth within two years before the survey. This is because the PNC data were collected from mothers who had given birth within two years of data collection, which may introduce some recall bias. Conversely, the findings of this study were lower than those reported in studies conducted across 33 and 17 sub-Saharan African (SSA) countries, a systematic review in Africa, and studies in nine SSA and Southeast Asian countries, which found rates of 64.2%, 75%, 79.1%, and 83%, respectively (16, 18, 51–53). This discrepancy may be due to the pooled proportion of dropouts in the current study, which included additional regions beyond SSA and Southeast Asia. Another factor could be the inclusion of more recent DHS survey data in the current study, whereas previous studies only included DHS data collected before 2020, potentially leading to a decrease in dropout rates in this study. Furthermore, discrepancies might result from the exclusion of PNC services delivered after 24 hours in the previous study. While mothers with at least one ANC visit are considered part of the continuum of care, previous studies that included only PNC within 24 hours may have reported higher dropout rates (16). Additionally, some studies incorporated the continuum of neonatal and child health care, unlike the current study, which may have contributed to higher dropout rates in those analyses (53).

The findings of this study are also lower than those reported in other studies conducted in Ethiopia, Nigeria, Ghana, Tanzania, the Lao People’s Democratic Republic, and India (17, 29, 54–66). This discrepancy may be attributed to differences in the composition of the study participants. The current study was based on the pooled prevalence from the DHS reports of multiple countries, which might reduce the overall proportion of dropouts. Another reason could be differences in the inclusion of outcome variables; previous studies included neonatal and infant postnatal care (PNC), unlike the current study (17, 66). The findings were also higher than those of previous studies conducted in Ethiopia and Cambodia (19, 37). This discrepancy may be attributed to differences in the operational definition of postnatal care (PNC) visits. The current study considered PNC within 48 hours postpartum, whereas previous studies assessed PNC within six weeks. This narrower time frame may have contributed to a higher dropout rate in the current study. Additionally, differences in health facility accessibility may explain the variation. Some prior studies were conducted in urban areas (37), where proximity to health facilities likely reduces dropout rates due to greater maternal care utilization and increased awareness of perinatal complications. These factors may account for the lower dropout rates observed in previous studies compared to the current one.

According to Table 3, regarding the factors associated with dropout from the maternal continuum of care in the present study, eleven variables (educational status, WHO region, media exposure, wealth quintile, parity, age of the respondent, getting permission to seek healthcare, distance to a health facility, timing of first ANC initiation, place of residence, and pregnancy intention) were found to be statistically significant. Women’s education was negatively associated with dropout from the maternal continuum of care. This finding aligns with studies conducted in sub-Saharan Africa, Nigeria, Egypt, Ethiopia, Nepal, Ghana, and the Lao People’s Democratic Republic, which show that as education levels increase, the likelihood of dropout decreases (17, 18, 35, 59, 61, 66–72). Higher education levels enable women to gain knowledge and awareness about the risks associated with discontinuing maternal health care services (35). Additionally, greater educational attainment may increase the demand for healthcare (73). Educated women tend to seek quality care and pay closer attention to their health to ensure better outcomes for themselves and their newborns.

Consistent with our findings, studies conducted in Sub-Saharan Africa (SSA), Egypt, and Ethiopia show that women with regular access to media have lower odds of dropping out from the continuum of care (CoC) (18, 35, 37, 59, 71, 74). Media are among the most important and powerful tools for disseminating information about maternal health. They can influence women’s knowledge, attitudes, and awareness regarding the risks associated with discontinuing maternal healthcare services. Similarly, the current study aligns with research from SSA and Ethiopia, which found that distance from health facilities is not a significant factor reducing the likelihood of dropout from CoC (51, 59, 70). This may be due to improved transportation accessibility. However, the use of maternal health services can be affected by the distance travelled from home to the healthcare facility (75, 76). Mothers living farther from health facilities have an increased chance of dropping out of maternal health services due to transportation challenges or other difficulties (61, 63, 77). Conversely, as the distance to healthcare facilities decreases and mothers live closer, they are more likely to receive maternal healthcare services (59, 63, 69, 70).

In the current study, difficulty getting permission to seek healthcare significantly increased the likelihood of dropout. This finding aligns with a study conducted in Ethiopia (70). Maternal care utilization was significantly associated with women acting as the primary decision-makers regarding the maternity continuum of care (51, 61, 71, 74). Women living in households led by females were more likely to use maternal health services (18). A woman’s ability to make decisions about her maternal healthcare is critical to completing all components of care (78).

Our findings indicate that, in a study conducted in Egypt, the age group between 20 and 35 years was associated with decreased odds of dropout from the continuum of care (CoC) (35). Consistent with our study, research in Ethiopia and sub-Saharan Africa (SSA) has shown that as age increases, dropout from CoC decreases (16, 18, 69). This trend may be explained by the fact that older maternal age is associated with greater knowledge and awareness of pregnancy-related complications and perinatal outcomes. Consequently, older mothers are less likely to drop out of care. In contrast, studies conducted in Ethiopia and Nigeria have reported that dropout rates increase with age (61, 79). This discrepancy might be due to younger women, particularly those under 20 years, experiencing greater fear of pregnancy-related outcomes.

Regarding parity, our findings align with a study from Ethiopia, which found that higher parity is associated with increased dropout from the continuum of care (63). This may be because nulliparous women are more concerned about pregnancy-related complications and thus more likely to seek care consistently. Conversely, women with multiple prior pregnancies often feel confident in managing their maternal health based on past experiences, which may reduce their likelihood of seeking healthcare services. However, our findings differ from those of another study in Ethiopia, which found that dropout rates decline with higher numbers of childbirths. (71). Multiparous women may have had more frequent contact with healthcare providers during previous pregnancies and are likely to be better informed about the potential risks of dropping out of the continuum of care (CoC). Additionally, multiparous mothers may face a higher risk of perinatal complications (80). For this reason, dropout rates might decrease with higher parity. Studies conducted in Nigeria, Egypt, Tanzania, Malawi, and Ethiopia support our findings, showing that women in lower wealth quintiles have higher odds of dropping out from the CoC (17, 35, 61–63, 68). Women who live in urban areas are more likely to benefit from better healthcare infrastructure, closer proximity to healthcare services, and greater financial means to cover healthcare costs. Furthermore, women in higher wealth index groups tend to be more educated and have better awareness of perinatal complications (35, 70). The likelihood of a woman receiving maternal healthcare services decreases with lower socioeconomic status (17, 61, 68, 81). Along the maternal continuum of care, dropouts are significantly influenced by the interaction between wealth index and place of residence. Rural women are particularly disadvantaged due to barriers such as limited access to healthcare services, transportation difficulties, and financial constraints. These challenges frequently result in interruptions in accessing essential maternal health services (82). Consistent with findings from studies conducted in Ethiopia, Nigeria, and other Sub-Saharan African countries, living in rural areas is associated with an increased dropout rate (18, 61, 69, 74, 83). This may be attributed to a lack of awareness among mothers regarding the consequences of dropout, as mothers in rural areas often have limited access to formal education. These mothers are more likely to discontinue the service (59, 61). Additionally, infrastructure challenges and the long distance from home to health facilities may contribute to higher dropout rates (61, 63, 75, 77).

Consistent with studies conducted in Sub-Saharan Africa (SSA) and Ethiopia, women who initiated antenatal care (ANC) follow-up late were more likely to drop out of the maternal continuum of care (18, 58, 63). Late initiation of ANC misses the opportunity to discuss birth preparedness and complication readiness plans. Consequently, these women may be less aware of danger signs during pregnancy, delivery, and the postpartum period. For this reason, the current study found that late initiation of ANC was associated with a higher likelihood of dropout from the continuum of care (CoC). Targeted messages and policies may be necessary to reduce dropout rates among mothers of varying wealth statuses, ages, and parity (84, 85). Additionally, women who planned their pregnancies were less likely to drop out of CoC, a finding consistent with studies conducted in SSA and Ethiopia (37, 51, 71, 74). Unplanned pregnancies may lead to delayed ANC initiation due to a lack of psychological preparedness or concerns about disclosure, leaving pregnant women unaware of potential pregnancy complications.

Regarding the WHO regions involved in the study, the risk of dropout from the continuum of care was higher in Sub-Saharan Africa (SSA), Central Asia, Southeast Asia (SSEA), Oceania, and Latin America and the Caribbean compared to the North Africa/West Asia/Europe region. In several SSEA countries, most countries in SSA, Oceania, Latin America, and the Caribbean, dropout rates were higher than the pooled average. This disparity may be attributed to the significant progress made by countries in the North Africa/West Asia/Europe region in achieving the Millennium Development Goals and their notable advancements toward the Sustainable Development Goals (SDGs) (38, 39). Countries in this region are classified as upper-middle-income economies (31), which likely contributes to the reduced dropout rates. It is also important to note that the study included only one country from each of Oceania, Central Asia, Latin America, and the Caribbean. Therefore, the reported proportions may not accurately represent the actual figures for these regions.

### Strengths and limitations of the study

DHS data from 41 LMICs provide a large, nationally representative, and standardized dataset that enables robust cross-country comparisons, enhances the generalizability of findings, and supports evidence-based policy formulation at both national and global levels. The use of comprehensive data on standard maternal health indicators to identify factors associated with dropout from the maternal continuum of care, along with the sampling design, made the analysis the main strength of this study. These advantages stemmed from utilizing extensive, nationally representative data from 41 low- and middle-income countries (LMICs), employing multilevel statistical analysis, and achieving a very high response rate in the survey interviews. However, using secondary data limited our ability to examine the effects of various factors that might influence maternal behavior, such as the mother’s knowledge of CoC dropout, perceptions of care quality, and cultural practices. Excluding these variables could have compromised cross-country comparisons. Furthermore, the main CoC components were measured through self-reporting based on women’s recall, which may have introduced recall bias. This bias is plausible because all information regarding CoC dropout relied on maternal memory of the services received. Additionally, some important variables, such as maternal occupation, husband’s education and occupation, and awareness of pregnancy complications, were excluded due to incomplete data, which may have affected the findings.

## Conclusions and Recommendations

The findings of this study indicate that a larger proportion of women in the study area discontinued the Continuum of Care (CoC). The global target of each component of maternal care (ANC4+, SBA, and PNC) is expected to exceed 80% (2). Based on this evidence, the acceptable dropout rate from the CoC should be less than 20%. These findings indicate that LMICs require more comprehensive efforts to achieve maternal and child health-related goals (SDG 3) by 2030. Distance to a health facility, getting permission, delayed initiation of the first antenatal care (ANC) visit, high parity, rural residency, and being from Sub-Saharan Africa (SSA), Southeast Asia (SSEA), Oceania, and Latin America and the Caribbean are factors positively associated with the outcome of interest. Conversely, women’s educational attainment, media exposure, wealth index, and maternal age are factors negatively associated with dropout from the maternal continuum of care.

Several factors significantly contribute to dropout from the continuum of maternal care. Improving women’s education, reducing adolescent pregnancies (before age 20), enhancing media exposure, increasing access to health facilities in remote societies, and increasing women’s decision-making power were identified as key modifiable factors to reduce dropout rates in the maternity continuum of care. WHO regions such as Sub-Saharan Africa (SSA), Southeast Asia (SSEA), Oceania, Latin America, and the Caribbean require urgent attention to reduce dropout rates. Significant efforts are needed to meet the maternal and child health Sustainable Development Goals (SDGs) by 2030. The proportion of dropouts was significantly higher in countries including Ethiopia, the Democratic Republic of Congo, Guinea, Mozambique, Mauritania, Burundi, Haiti, Madagascar, Timor-Leste, Bangladesh, Uganda, Papua New Guinea, Mali, Nigeria, Rwanda, Tanzania, Benin, and Côte d’Ivoire. Those proportions exceed the pooled average. To meet the SDG targets by 2030 in these countries, immediate and focused efforts are required.

Different stakeholders are expected to undertake tasks to improve these factors. The WHO and the health authorities of these 41 low- and middle-income countries, particularly those with dropout rates exceeding the overall average, need to promptly implement measures to decrease dropout rates to meet the Sustainable Development Goals (SDG 3.1 and 3.2). In particular, countries from Sub-Saharan Africa (SSA), South and Southeast Asia (SSEA), Oceania, and the Latin America and the Caribbean regions are strongly encouraged to address dropout rates. Initiating antenatal care (ANC) before the first trimester requires a short time but significantly contributes to reducing dropouts. Health authorities and professionals should emphasize the importance of beginning the first ANC visit before 12 weeks of gestation. Healthcare providers and community health workers must collaborate to strengthen linkages between antenatal care, skilled delivery, and postnatal follow-up. Increasing community access to media and providing health education to mothers and influential community leaders through targeted campaigns is an effective strategy to raise knowledge and awareness about the risks associated with dropping out of maternity care. For researchers, these findings serve as a valuable source of information for conducting further studies in this field, including studies with psychological and behavioral factors, different contexts, and the impact of dropout from the continuum of care on birth outcomes.

## Data Availability

All data produced in the present work are contained in the manuscript.

## Acronyms

AOR: Adjusted Odd Ratio
ANC: Antenatal Care
DHS: Demographic and Health Survey
CI: Confidence Interval
CoC: Continuum of maternity Care
ICC: Intra-class Correlation Coefficient
LMICs: Low- and Middle-Income Countries
MOR: Median Odds Ratio
PCV: Proportional Change in Variance
PNC: Postnatal Care
SBA: Skilled Birth Attendant
SDG: Sustainable Development Goals
SSA: Sub-Saharan Africa
SSEA: South and Southeast Asia
VIF: Variance Inflation Factor;
WHO: World Health Organization

## Acknowledgments

The authors wish to express our gratitude to the DHS program for allowing us to access and use the data online.

## Authors’ contributions

MDB led the conceptualization, data cleaning, formal analysis, methodology development, software use, validation, visualization, initial manuscript drafting, and subsequent revisions and editing. AM, MS, TDK, and MG assisted with data cleaning, formal analysis, methodology, software use, manuscript drafting, reviewing, and editing. All authors reviewed and gave their approval for the final manuscript version.

## Declarations

### Ethical approval

The Institutional Review Board and the health service ethical review committees of the included countries approved the data collection for all DHS surveys. The benefits and risks of the surveys were explained to the participants. Before administering the Women’s Questionnaire, eligible respondents provided written informed consent. Participation in the survey was entirely voluntary. The names and identification numbers of respondents were excluded from the final datasets. Ethical approval was not required for using existing public domain datasets because these data are freely accessible online with all personal identifiers removed. To obtain the data, we submitted an online request, received an authentication letter, and obtained explicit permission from the DHS program.

### Patient and public involvement

This study examined anonymized, publicly available data from the DHS. No patients or members of the public directly designed, carried out, or reported this study.

### Consent for publication

Not applicable.

